# COVID Seq as Laboratory Developed Test (LDT) for diagnosis of SARS-CoV-2 Variants of Concern (VOC)

**DOI:** 10.1101/2022.11.11.22282032

**Authors:** Rob E. Carpenter, Vaibhav K. Tamrakar, Sadia Almas, Emily Brown, Rahul Sharma

**Affiliations:** Advanta Genetics, 10935 CR 159, Tyler, Texas 75703, USA; University of Texas at Tyler, 3900 University Boulevard, Tyler, Texas 75799, USA; ICMR-National Institute of Research in Tribal Health, Jabalpur, MP 482003, INDIA; RetroBioTech LLC, 838 Dalmalley Ln, Coppell, TX 75019, USA; Scienetix, 10935 CR 159, Tyler, Texas 75703, USA

**Author notes:** **Corresponding author:** Rahul Sharma PhD, Advanta Genetics, 10935 CR 159 Tyler, Texas 75703, USA.

**Keywords:** SARS-CoV-2, Variants of Concern, Laboratory Developed Test (LDT), CLIA, Next Generation Sequencing

## Abstract

Rapid classification and detection of SARS-CoV-2 variants have been critical in comprehending the virus’s transmission dynamics. Clinical manifestation of the infection is influenced by comorbidities such as age, immune status, diabetes, and the infecting variant. Thus, clinical management may differ for new variants. For example, some monoclonal antibody treatments are variant-specific. Yet, an FDA-approved test for detecting the SARS-CoV-2 variant is unavailable. A laboratory-developed test (LDT) remains a viable option for reporting the infecting variant for clinical intervention or epidemiological purposes. Accordingly, we have validated the Illumina COVID-Seq assay as an LDT according to the guidelines prescribed by the College of American Pathologists (CAP) and Clinical Laboratory Improvement Amendments (CLIA). The limit of detection (LOD) of this test is Ct<30 (∼15 viral copies) and >200X genomic coverage, and the test is 100% specific in the detection of existing variants. The test demonstrated 100% precision in inter-day, intra-day, and intra-laboratory reproducibility studies. It is also 100% accurate, defined by reference strain testing and split sample testing with other CLIA laboratories. Advanta Genetics LDT COVID Seq has been reviewed by CAP inspectors and is under review by FDA for Emergency Use Authorization.

## 1. INTRODUCTION

Severe Acute Respiratory Syndrome (SARS) SARS-CoV-2 is the etiological agent of COVID-19, which is associated with mild respiratory symptoms in most infections. However, for patients with underlying medical conditions, comorbidities, and advanced age, COVID-19 may lead to severe illness. The primary route of SARS-CoV-2 transmission between humans is the respiratory route, including droplets of saliva or discharge from infected patients. Diagnosis of COVID-19 relies on detecting SARS-CoV-2 viral RNA from a nasopharyngeal or oropharyngeal specimen [1]. However, the rapid emergence of several variants with higher virulence and infectivity has provoked repeat waves of the deadly pandemic in many countries and raised anxieties about vaccine efficacy and diagnostic accuracy [2]. Rapid classification and tracking of emerging variants are critical for understanding the transmission dynamics of this disease and developing strategies for severing the transmission chain. Next-Generation Sequencing (NGS) remains the tool of choice for whole-genome analysis and deciphering new mutations [3]. Within a relatively short period, SARS-CoV-2 has acquired several mutations resulting in different virus variants. In December 2020, the United Kingdom reported a SARS-CoV-2 variant of concern (VOC), lineage B.1.1.7, detected in over 30 countries and is more efficiently transmitted than other SARS-CoV-2 variants. Thus, the pandemic strikes in several phases of outbreaks in different parts of the world [4]. Currently, the virus continues to be a global agent of infection.

The highly mutagenic nature of SARS-CoV-2 assaulted many countries with second or third waves of the outbreak [5, 6]. Mutations with higher transmissibility, a more intense disease state, and less likely to respond to vaccines or treatments have been classified by the World Health Organization (WHO) as Variants of Concern. Recent epidemiological reports released by WHO indicated five VOCs: 1) B.1.1.7 (Alpha) in December 2020; 2) B.1.351 (Beta) in December 2020; 3) P.1 (Gamma) in January 2021; 4) B.1.617.2 (Delta) in December 2020, and 5) B.1.1.529 (Omicron) (https://www.who.int/en/activities/tracking-SARS-CoV-2-variants/). The receptor-binding domain (RBD) of coronavirus increases its capacity to strike in several outbreak phases in different parts of the world [7]. More recently, South Africa reported a new SARS-CoV-2 variant to the WHO.

Omicron (B.1.1.529) was first detected in specimens collected in Botswana and designated as the fifth VOC [8] (https://www.who.int/activities/tracking-SARS-CoV-2-variants). Several variant-specific treatment options have been approved by the U.S. Food and Drug Administration (FDA), including Bebtelovimab, a monoclonal antibody for the treatment of COVID-19 that retains activity against the omicron variant. However, a recent study shows that the effectiveness of mRNA vaccines is reduced against all three subvariants of omicron [9]. Several other studies have reported a substantial decrease in neutralizing antibody titers after vaccination against all coronavirus variants. [10, 11].

Reduced neutralizing activity against the B.1.1.7 (Alpha), B.1.35 (Beta), and P.1(Gamma) strains have been reported among the Pfizer-BioNTech vaccinated populations [12]. Another study investigated the neutralization of antibodies elicited by Novavax NVX-CoV2373, a protein subunit vaccine, and that of mRNA-1273 by Moderna against the California variant B.1.429 and B.1.351 pseudoviruses. The small-scale study of 63 volunteers similarly revealed the reduction in neutralization abilities of antibodies elicited by both vaccines. The most drastic reduction, up to a 9–14 times decrease in neutralization compared to D614G, was observed with B.1.351 pseudovirus, where the antibodies were 2–3 times less sensitive against the B.1.429 variant pseudovirus [13]. SARS-CoV-2 is likely to continue to evolve, and the next strain may have a strain-specific etiology requiring strain information for patient care.

In the present situation, most infections are attributed to a single sublineage. However, new lineages are likely to emerge and replace existing circulating lineages. Several PCR-based assays are available for the detection of the known variants. These assays are not designed to detect unknown infected variants [14]. Unlike the PCR-based test, this NGS-based assay can detect new variants as they emerge. And because lineage variance has potential implications for virulence and infectivity, validation of NGS assays that proactively identify mutagenic variants enables these test results to be used in clinical applications when warranted.

Furthermore, this Illumina COVID Seq assay is used for epidemiological surveillance globally. Still, the validation of the assay as a Laboratory Developed Test (LDT) is required to use variant information for clinical decision-making. For example, variant-specific monoclonal antibody therapies have been emphasized by the NIH COVID Treatment Guidelines Panel and the FDA, recommending against the use of bamlanivimab and etesevimab (administered together) and REGEN-COV (casirivimab and imdevimab) because of significantly reduced activity.

Consequently, we report the validation of the NGS-based test to identify the existing and emerging variants of SARS-CoV-2 [16]. This study has benchmarked the validation process for using the variant information in clinical management as required by CLIA. Although the Illumina COVID-Seq assay has been approved for emergency use authorization (EUA) for the diagnosis of COVID-19; the assay has not been approved for variant detection. We validated the Illumina COVID-Seq assay according to CLIA/CAP requirements for LDT, and the validation report has been submitted to the FDA for EUA, and reviewed by a team of CAP inspectors. Accordingly, the COVID-Seq assay is qualified to diagnose SARS-CoV-2 variants of infected individuals and can be deployed for monitoring the evolution of SARS-CoV-2 variants in decentralized clinical laboratory settings.

## 2. MATERIALS AND METHODS

The workflow consists of the following procedures: RNA extraction, cDNA synthesis, target amplification, library preparation, library pooling, sequencing, and analysis. Validation was performed to achieve a high degree of accuracy and precision. Additional studies were performed to test the effect of interference substances and sample stability.

### 2.1 Reference strains of SARS-CoV-2 variants

We used three reference strains of SARS-CoV-2, Omicron, Delta, and Wuhan. Complete genome synthetic RNAs of these strains were obtained from BEI Resources.

### 2.2 Clinical specimens

De-identified sample remnants from nasopharyngeal swabs were collected from the patients who tested positive for SARS-CoV-2 PCR with RT-PCR at Advanta Genetics (https://aalabs.com/) in Tyler, Texas. Samples were stored at −80°C until RNA extraction. The study was exempted by IRB (Institutional Review Board) because only de-identified samples were used.

### 2.3 RNA Extraction

Total RNA was extracted using the Roche MagNA Pure 96 System and Viral RNA Small Volume Kits per the manufacturer’s (Port Scientific Inc. QC J3G 4S5 Canada) instructions. Isolated RNA was frozen at −80°C until the library preparation.

### 2.4 Library Preparation and Sequencing

The libraries were prepared using the Illumina COVID-Seq protocol (Illumina Inc, USA). Briefly, total RNA was primed with random hexamers, and first-strand cDNA was synthesized using reverse transcriptase. The SARS-CoV-2 genome was amplified using the two sets of primers (COVID-Seq Primer Pool-1 & 2) provided by Illumina, but the primer sequences have not been disclosed by the manufacturers. Primers are not mutation specific but designed to amplify the entire genome. PCR amplicons were tagmented using the EBLTS (Enrichment BLT), which is a process that fragments and tags the PCR amplicons with adapter sequences. Adaptor ligated amplicons were further amplified using the distinct pre-paired 10 base pair Index 1 (i7) adapters and Index 2 (i5) adapters (IDT for Illumina-PCR Indexes Set 1) for each sample. The individual library was quantified using a Qubit 2.0 fluorometer (Invitrogen, Inc.) and pooled in equimolar concentration instead of equal volume as recommended by Illumina. This additional step allowed us to achieve uniform coverage of all the libraries in the pool and efficient use of a low throughput sequencing instrument (MiniSeq®). A COVID-Seq positive control (Wuhan-Hu-1) and one no template control (NTC) were processed with each batch of libraries. The final library pool was again quantified using a Qubit 2.0 fluorometer (Invitrogen, Inc.) and a PCR-based library quantification kit (Scienetix, USA). The final library pool was diluted to a 2 pM loading concentration. Dual indexed paired-end sequencing with 75bp read length was carried out using the HO flow cell (150 cycles) on the Illumina MiniSeq® instrument.

### 2.5 NGS Data Analysis

Illumina Basespace (https://basespace.illumina.com) bioinformatics pipeline was used for sequencing QC, FASTQ generation, genome assembly, and identification of SARS-CoV-2 variants. Briefly, the raw FASTQ files were trimmed and checked for quality (Q>30) using the FASTQ-QC application within the Basespace. QC passed FASTQ files were aligned against the SARS-CoV-2 reference genome (NCBI Reference Sequence NC_045512.2) using Bio-IT Processor (Version: 0×04261818). Then, DRAGEN COVID Lineage (Version: 3.5.4) application in Bbasespace was used for SARS-CoV-2 variant determination and generating a single consensus FASTA file. Finally, single consensus FASTA was also analyzed for lineage assignment using the web version of Phylogenetic Assignment of Named Global Outbreak Lineages (PANGOLIN) software (https://pangolin.cog-uk.io). Only the consensus variants identified by both applications were used for further analysis.

### 2.6 Strain typing of SARS-CoV-2 in the East Texas region

We have applied the SARS-CoV-2 variant detection workflow established in this study for strain typing of SARS-CoV-2 in the East Texas USA region over the course of the pandemic. Representative samples collected at various po time during the pandemic (Aug 2020, July 2021, Dec 2021, April 2022, July 2022, and Sept 2022) were sequenced and analyzed for the circulating variants in the region of interest.

## 3. RESULTS

The SARS-CoV-2 sequencing test was validated as LDT according to the guidelines prescribed by the CAP and mandated by CLIA. Briefly, the limit of detection (LOD), analytical accuracy, precision, and sample stability was established. The effect of carryover and interference substances was also investigated.

### 3.1 Analytical Sensitivity/Limit of Detection

This assay is not intended to diagnose the SARS-CoV-2 infection but is meant for discovering the SARS-CoV-2 variant from a patient previously diagnosed with a high level (Ct<30) of SARS-CoV-2 infection. LOD of this assay was determined for two variables needed for accurate results: 1) the lowest amount of input genomic material and 2) minimum genomic coverage. For genomic material LOD, serial dilutions of an Omicron reference variant were sequenced in triplicates, and the lowest input concentration resulting in the correct variant detection is identified as LOD (Table-1). LOD for the test is defined as Ct<30 (∼15 copies/ul of RNA). LOD was further verified by sequencing 23 samples with RNA input close to LOD (PCR Ct value ∼30±2) and obtained 200X-1000X coverage; variants for all the 23 samples were identified correctly. We have also analyzed the 26 additional samples from patients found positive during September 2022—all the samples have Ct<30. We sequenced the 26 samples, including 10-fold and 100-fold dilutions of three representative samples. We were able to identify the variant in all 26 samples and endorse the application of the assay on current circulating strains post-vaccination. However, the application of this test in asymptomatic or very low viral load (<150 viral genome/ul) remains the limitation of the assay.

**Table-1:** Limit of Detection: Four serial dilutions of Omicron strain were sequenced in duplicate, and the lowest viral RNA input, which resulted in accurate variant detection, was accepted as LOD for the test.

| Pango Lineage | Sample ID | Relative Copy/ul | ACSQ Observed Lineage |  |  |  |  |  |
| --- | --- | --- | --- | --- | --- | --- | --- | --- |
| | | | PCR Ct Value | | Median Coverage | Coverage $\geq 30x$ | Pango Lineage | WHO label |
|  |  |  | N1 | N2 |  |  |  |  |
| **BA.1** | ACSQ4-19 | 1.48E+04 | 26.16 | 26.37 | 663 | 96.79% | BA.1 | Omicron |
|  | ACSQ4-19 (1:10) | 1.48E+03 | 29.87 | 30.15 | 690 | 95.18% | BA.1 | Omicron |
|  | ACSQ4-19 (1:100) | 1.48E+02 | 33.24 | 33.54 | 504 | 94.46% | BA.1 | Omicron |
|  | ACSQ4-19 (1:1000) | 1.46E+01 | 36.54 | 35.47 | 586 | 91.29% | BA.1 | Omicron |
| **BA.1** | ACSQ4-19 | 1.48E+04 | 26.16 | 26.37 | 533 | 95.87% | BA.1 | Omicron |
|  | ACSQ4-19 (1:10) | 1.48E+03 | 29.87 | 30.15 | 489 | 94.89% | BA.1 | Omicron |
|  | ACSQ4-19 (1:100) | 1.48E+02 | 33.24 | 33.54 | 745 | 94.97% | BA.1 | Omicron |
|  | ACSQ4-19 (1:1000) | 1.46E+01 | 36.54 | 35.47 | 736 | 93.61% | BA.1 | Omicron |
| **BA.4.6** | ACSQ9-1A | 1.48E+03 | 29.83 | 28.36 | 1597 | 94.25% | BA.4.6 | Omicron |
|  | ACSQ9-1A (1:10) | 1.48E+02 | 33.16 | 31.69 | 330 | 82.33% | BA.4.6 | Omicron |
|  | ACSQ9-1A (1:100) | 1.46E+01 | 36.49 | 35.02 |  |  |  | Not detected |
| **BA.5.2.1** | ACSQ9-4A | 1.58E+03 | 27.4 | 26.17 | 2300 | 96.89% | BA.5.2.1 | Omicron |
|  | ACSQ9-4A (1:10) | 1.58E+02 | 30.73 | 29.5 | 1635 | 92.93% | BA.5.2.1 | Omicron |
|  | ACSQ9-4A (1:100) | 1.58E+01 | 34.06 | 32.83 | 514 | 88.59% | BA.5.2.1 | Omicron |
| **BA.2.3** | ACSQ9-23 | 1.48E+04 | 26.65 | 23.71 | 3355 | 99.46% | BA.2.3 | Omicron |
|  | ACSQ9-23 (1:10) | 1.48E+03 | 29.98 | 27.04 | 2798 | 98.52% | BA.2.3 | Omicron |
|  | ACSQ9-23 (1:100) | 1.48E+02 | 33.31 | 30.37 | 2051 | 95.68% | BA.2.3 | Omicron |

To determine the LOD regarding genomic coverage, we computed the depth of coverage (X times) and percent genome coverage for all tested samples. The lowest genomic coverage of >200X (Depth) and 90% genome coverage is required for successful detection variant detection (Figure-1). Importantly, all 164/164 (100%) observations with a minimum of 90% genome coverage at a minimum of 200X resulted in the correct variant call after the analysis.

**Figure-1:**
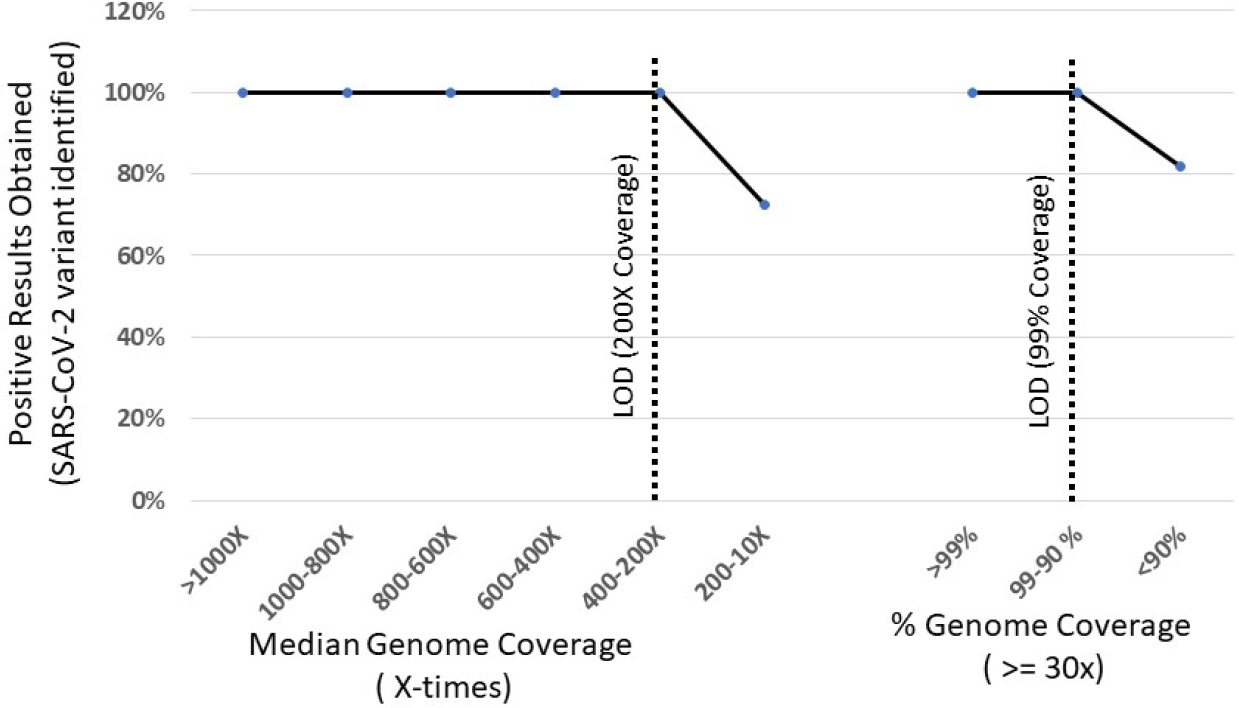
Limit of Detection (LOD): Median genomic coverage (X-times) and minimum % length of the genome covered >30X times were computed, and the minimum coverage required for obtaining the accurate SARS CoV-02 lineage was defined as LOD.

### 3.2 Analytical Accuracy

Accuracy is a determination of the amount of systematic error in the system. The analytical specificity of this assay is determined by re-sequencing the already sequenced reference strains of the SARS-CoV-2 virus and the alignment of the resulting FASTQ files to the available reference genome sequence using the BaseSpace (Illumina) tool. We tested 3 known SARS-CoV-2 variants; Wuhan-Hu-1, B.1.617.2 (Delta), and B.1.1.529 (Omicron) in triplicates, and all the variants were identified correctly as expected. NGS does not use analyte (i.e., SARS-CoV-2 variant) specific reagents to determine the correct variant but uses whole genome analysis for discerning the variants. Therefore, the specificity of the variant detection is considered 100% (Table 2).

**Table-2.** Accuracy of the test is determined by sequencing 3 reference strains of the SARS-CoV-2 virus

| Sample ID | Reference Lineage |  |  | Detected Lineage |  |  | Concordance |
| --- | --- | --- | --- | --- | --- | --- | --- |
|  | Median Coverage | Pango Lineage | WHO label | Median Coverage | Pango Lineage | WHO label |  |
| Wuhan (1500cp/ul) | NA | A1 | Non-VOC | 1167 | A1 | Non-VOC | YES |
| Wuhan (150cp/ul) | NA | A1 | Non-VOC | 2289 | A1 | Non-VOC | YES |
| Omicron (1500cp/ul) | NA | B.1.1.529 | Omicron | 1372 | B.1.1.529 | Omicron | YES |
| Omicron (150cp/ul) | NA | B.1.1.529 | Omicron | 829 | B.1.1.529 | Omicron | YES |
| Delta (1500cp/ul) | NA | B.1.617.2 | Delta | 663 | B.1.617.2 | Delta | YES |
| Delta (150cp/ul) | NA | B.1.617.2 | Delta | 533 | B.1.617.2 | Delta | YES |

Considering the limited availability of reference strains, we also re-sequenced the 6 samples already sequenced by another reference laboratory (Fulgent Genetics) at extremely high coverage (>30,000X), and variant identities were compared between two observations. A total of 6 samples were sequenced at Fulgent Genetic and Advanta Genetics. All 6 samples were identified to carry identical variants by both laboratories implicating 100% accuracy in inter-laboratory testing (Table-3). The average sequencing coverage at Fulgent Genetics is 34560.5X compared to 174.3X at Advanta. Interestingly, variants of 3 samples sequenced at >50,000 coverage were correctly identified by only 200X coverage. With pre-pooling quantification, we achieved higher sequencing efficiency without compromising the test accuracy. Such higher efficiency is critical for the cost-effective application of this test in limited-resourced and de-centralized laboratory settings.

**Table-3:** Comparative genome sequencing and variant calling results obtained from two different laboratories.

| Sample ID | Fulgent Genetics* |  |  | Advanta Genetics |  |  | Concordance |
| --- | --- | --- | --- | --- | --- | --- | --- |
|  | Median Coverage | Pango Lineage | WHO label | Median Coverage | Pango Lineage | WHO label |  |
| ACSQ1-2 | 77387.8 | AY.4 | Delta | 214 | AY.25 | Delta | YES |
| ACSQ1-3 | 341.6 | AY.3 | Delta | 228 | AY.3 | Delta | YES |
| ACSQ1-4 | 54823.9 | AY.3 | Delta | 222 | AY.3 | Delta | YES |
| ACSQ1-5 | 2860.5 | AY.4 | Delta | 218 | AY.3 | Delta | YES |
| ACSQ1-6 | 69088.9 | B.1.617.2 | Delta | 258 | B.1.617.2 | Delta | YES |
| ACSQ1-7 | 2860 | AY.3 | Delta | 106 | AY.3 | Delta | YES |

\*Variant from AY4 to AY 25 were reported as AY4 in the sequencing results obtained from the Fulgent Genetics

### 3.3 Precision

The precision of a measurement system, related to reproducibility and repeatability, is the degree to which repeated measurements under unchanged conditions show the same results. Inter-day precision is determined by sequencing 9 samples of known genomic variants over three days. Nine samples were tested in three rounds of library preparations, sequencing, and data analysis. The identity of the variant detected across the three runs was compared. All 9 samples were identified correctly across the three sequencing instances implicating 100% precision across the 27 observations.

Inter and intra-day precision was determined by testing 6 clinical samples (near LOD) in triplicate during three rounds of library preparations, sequencing, and data analysis. The identity of the variant detected across the three runs was compared. Three samples that failed the pre-defined QC (library yield, reads, coverage, etc.) were excluded from the precision. Over three days, the remaining 51/54 observations were in 100% concordance for triplicate testing. All 6 samples were identified as the same variant in triplicate testing within a single batch implicating 100% intra-run precision (Supplementary-2). Likewise, the same samples resulted in identical variants when tested in three distinct batches of library preparation, sequencing, and data analysis (Supplementary-3). Thus, inter-day precision was also determined as 100%.

### 3.4 Stability Study

The stability of clinical samples at different temperatures was tested to simulate the temperature conditions during transportation. Samples identified as Omicron (n=3) and Delta (n=1) were placed at 4 different temperatures [Freezer (−20□C); Refrigerator (2-8□C); Room Temp (∼25□C); Elevated Temp (∼50□C)] to mimic the possible environmental conditions during the transportation. Samples were left for up to 7 days under these temperature conditions. Samples were retrieved at 24 hours, 3 days, and 7 days intervals, and RNA was extracted and stored at −80°C. RNA from all the samples in the stability study was tested in a single library preparation and sequencing batch. We were able to sequence and identify the SARS-CoV-2 variant of the samples kept at 20°C, 2-8°C, ∼25°C, and ∼50°C for 24 hours and 3 days. However, samples placed in elevated temperature conditions resulted in low-quality sequencing data, which did not result in variant detection. Overall, samples kept at an elevated temperature (∼50°C) over 3 days were unsuitable for variant detection by whole-genome sequencing (WGS). All the samples used for the stability study were of viral load close to LOD.

### 3.5 Freeze-thaw stability study

Extracted RNA was subjected to 2 and 3 freeze-thaw cycles, and RNA was processed in single Library preparation. RNA sample after 2 freeze-thaw cycles fail the pre-defined sequencing QC and could not be used for variant detection. Although sample or RNA storage conditions are unlikely to change the SARS-CoV-2 variant, >3 days of storage at high temperature (∼50°C) may cause the SARS-CoV-2 variant testing to fail or result inconclusive because of compromised data quality. Likewise, >2 freeze-thaw cycles for RNA also compromised the sequencing data quality. Thus, samples for SARS-CoV-2 variant detection should be kept at 4°C for 7 days and stored at −20°C for the long term (Table-4).

**Table-4:**
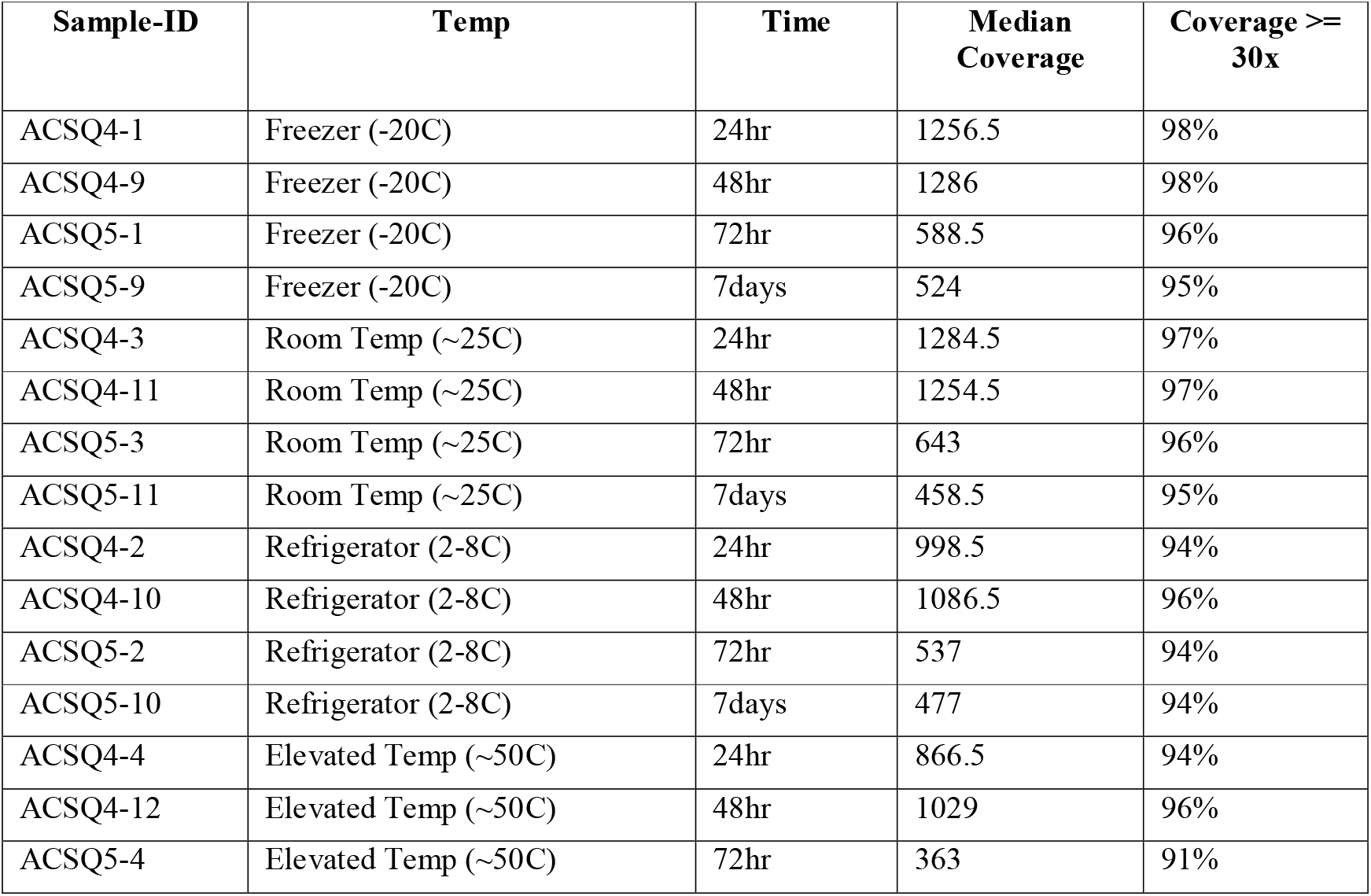
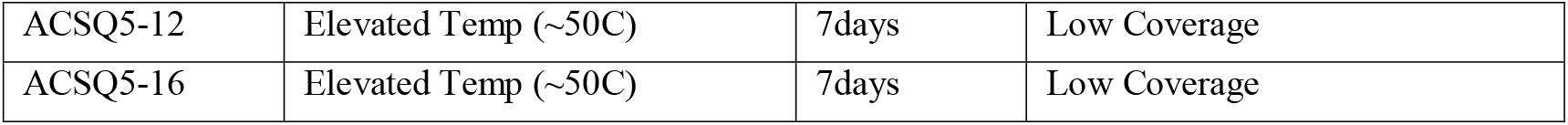
Samples stored in simulated environmental conditions mimicking the possible transportation and storage temperature were sequenced to identify acceptable sample storage conditions.

| Sample-ID | Temp | Time | Median Coverage | Coverage >= 30x |
| --- | --- | --- | --- | --- |
| ACSQ4-1 | Freezer (-20C) | 24hr | 1256.5 | 98% |
| ACSQ4-9 | Freezer (-20C) | 48hr | 1286 | 98% |
| ACSQ5-1 | Freezer (-20C) | 72hr | 588.5 | 96% |
| ACSQ5-9 | Freezer (-20C) | 7days | 524 | 95% |
| ACSQ4-3 | Room Temp (~25C) | 24hr | 1284.5 | 97% |
| ACSQ4-11 | Room Temp (~25C) | 48hr | 1254.5 | 97% |
| ACSQ5-3 | Room Temp (~25C) | 72hr | 643 | 96% |
| ACSQ5-11 | Room Temp (~25C) | 7days | 458.5 | 95% |
| ACSQ4-2 | Refrigerator (2-8C) | 24hr | 998.5 | 94% |
| ACSQ4-10 | Refrigerator (2-8C) | 48hr | 1086.5 | 96% |
| ACSQ5-2 | Refrigerator (2-8C) | 72hr | 537 | 94% |
| ACSQ5-10 | Refrigerator (2-8C) | 7days | 477 | 94% |
| ACSQ4-4 | Elevated Temp (~50C) | 24hr | 866.5 | 94% |
| ACSQ4-12 | Elevated Temp (~50C) | 48hr | 1029 | 96% |
| ACSQ5-4 | Elevated Temp (~50C) | 72hr | 363 | 91% |

|  |  |  |  |
| --- | --- | --- | --- |
| ACSQ5-12 | Elevated Temp (~50C) | 7days | Low Coverage |
| ACSQ5-16 | Elevated Temp (~50C) | 7days | Low Coverage |

### 3.6 Role of interference substances

Clinical specimens may contain biological or non-biological substances which may interfere with the testing process. We spiked the commonly used nasal sprays into the clinical specimen and tested the sample with and without the external substance. None of the tested substances altered the results or compromised the data quality (Data not shown).

### 3.7 Epidemiological survey of East Texas, US

We have also applied Advanta Genetics LDT COVID-Seq to investigate the evolution of SARS-CoV-2 in the East Texas region during the pandemic. We identified a greater genomic diversity in early pandemics before identifying variants of Concern. We identified the SARS-CoV-2 variant (B.1, B.1.126, B.1.2, B.1.234, B.1.243, B.1.564, B.1.574, B.1.602) among the samples collected in July 2020. All of these variants were categorized as non-VOC by the WHO. Diverse non-VOC strains were initially replaced by the Delta variant (100%) in July-Aug 2021. Omicron (58%) and Delta (42%) variants were co-circulating during Dec 2022; Delta was completely replaced by the Omicron variant by December 2021. Omicron BA.2 (79%) was the dominant variant during April 2022, which was again replaced by BA.5 (78%) in September 2022. (Figure-2). Thus, continuous monitoring is warranted to keep the pandemic from returning to the scale seen earlier by identifying the vaccine escape or target dropout in diagnostic testing. All the SARS-CoV-2 whole-genome sequences generated in this study were submitted to GISAID (https://www.gisaid.org) database (Supplementary Table-1), [17].

**Figure-2:**
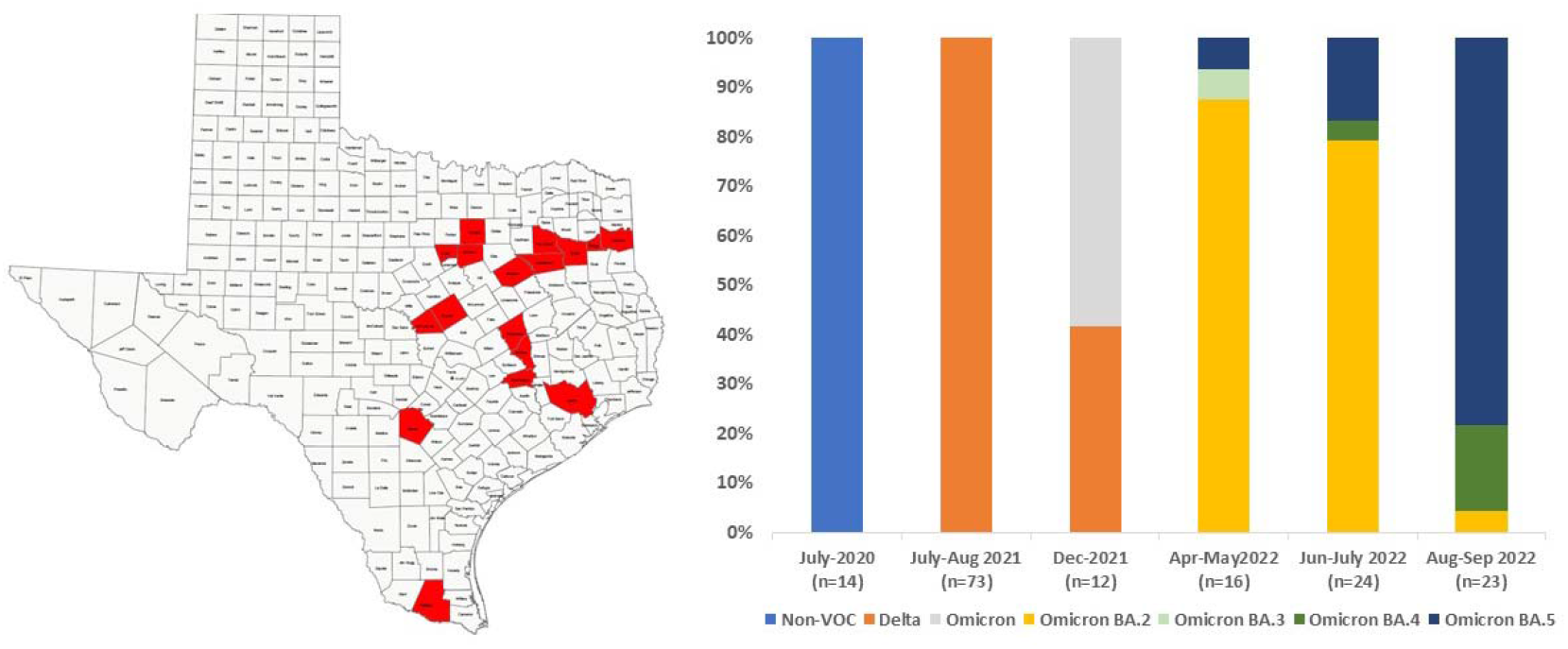
Evolution of SARS-CoV-2 variants in East Texas over the course of the Pandemic

## 4. DISCUSSION

WHO has been classifying the SARS-CoV-2 variant into various categories according to their possible clinical implication and public health concern. Several technologies have been adopted for SARS-CoV-2 variant detection, but NGS remains the gold standard because of comprehensive genomic analysis [18, 19]. In June 2020, the US FDA granted EUA for Illumina’s NGS test for COVID-19 diagnosis. However, the test has not been widely adopted for diagnosis because RT-PCR is much cheaper and easy to implement in the unprecedented need for SARS-CoV-2 testing. Although RT-PCR remains the method of choice for routine diagnosis, Illumina COVID-Seq protocol has been instrumental in outbreak investigation and surveillance throughout the pandemic [20]. Several laboratories worldwide use WGS for high throughput surveillance communicated by health organizations [21]. The Delta variant has been associated with greater transmissibility and higher viral RNA loads in both unvaccinated and fully vaccinated individuals [22]. WGS has also identified the potential compromise vaccine effectiveness against the Omicron variant [23]. The emergence of SARS-CoV-2 variants with significantly different clinical implications accentuates the need for variant detection, especially for immunocompromised patients. Additionally, because some monoclonal antibody treatments are variant-specific, timely identification of the infecting SARS-CoV-2 variant may influence decision-making and treatment. Currently, there is no FDA-approved SARS-CoV-2 variant detection test for diagnosing individual patients. Thus, LDT remains the only viable option to leverage NGS methods for SARS-CoV-2 variant diagnosis. This virus is predicted to mutate continuously, and the evolution of variants with significantly different clinical interventions cannot be ruled out [5].

This study established the WGS workflow for detecting SARS-CoV-2 variants according to CLIA guidelines for LDTs. The importance of reference materials for the validation and QC of wet-lab and dry-lab WGS processes is well established [24]. However, unlike human genomics [25], there is no well-established resource of reference materials for the validation of such genetic variant detection tests. Therefore, we obtained three reference strains (Wuhan, delta, and Omicron) of SARS CoV-2 for accuracy study, and all three variants were correctly identified in repeated testing. The Wuhan strain of SARS-CoV-2 has been accepted as the reference strain [26]. Therefore, all the sequences generated during this study were aligned against the Wuhan strain genome [26]. Although more than a million SARS-CoV-2 genomes have been sequenced [27], a limited number of well-characterized reference genomic materials are available. To overcome this limitation, we re-sequenced 6 samples already sequenced by another laboratory (Fulgent Genomics), and the sequencing results were in 100% concordance.

Interestingly, 3 of these samples were sequenced at >50,000X coverage by Fulgent Genetics, and the same samples were sequenced at 200-300X coverage at Advanta Genetics. Results from both sequencings were in 100% concordance, suggesting that such high sequencing depth is unnecessary for routine variant detection. The introduction of pre-pooling quantification and equimolar pooling enabled us to achieve uniform distribution of sequencing reads across the samples in the pool, resulting in more efficient sequencing. This approach particularly important in de-centralized reference laboratories which do not have access to high throughput instruments such as Illumina HighSeq or NovSeq. We were able to sequence up to ∼30 samples in a single MiniSeq run, reducing the cost of sequencing (excluding library preparation) to ∼ $30/sample. The genome sequences available from public databases may have been generated using different sequencing chemistries or platforms, which might yield different error rates; therefore, the inter-laboratory study was of particular interest because the reference laboratory used a different instrument for sequencing. Overall, we achieved high accuracy, reproducibility, repeatability, diagnostic (variant detection) sensitivity, and specificity of 100%, which exceeds the 90% threshold for LDT performance parameters per CLIA requirements.

These findings agree with other reports of 93% to 100% accuracy in WGS identification and subtyping for other pathogens [28, 29]. We determined the LOD as 90% genome sequenced at >30X depth and >200X median depth of coverage. The LOD study did not consider coverage for individual single nucleotide polymorphisms (SNP) because an SNP combination determines the SARS-CoV-2 genomic variant. LOD, in terms of minimum genomic copies, was established at ∼15 copies/μL going into the sequencing reaction. This assay can identify the genomics variant from the lower viral RNA input, but this is the lowest input tested during this validation. Vaccination has reduced hospitalization and deaths in COVID cases, but the viral load (10E+05 to 10E+08 genomic copies/ml) in breakthrough cases remains high enough for detection by this assay [15]. Interestingly, only infectious viral load (VL) was lower in fully vaccinated Omicron BA.1-infected individuals compared to vaccinated Delta-infected individuals, indicating variant-specific response to the vaccines. A reduced infectious VL) was observed only in boosted but not fully vaccinated individuals compared to unvaccinated individuals [15]. Still, genomic copies/ml of the sample remain very high (∼million genomic copies/ml), significantly above the LOD of this test, implicating that test will be useful in a post-vaccination era. We found some of the CLIA-defined LDT performance criteria difficult to apply. For example, CLIA would allow for up to 10% of base calls to be incorrect for accuracy determination, which, in the case of the ∼30,000 bp SARS-CoV-2 genome, would mean ∼3,000 inaccurate bases, which could lead to false variant detection. We accepted a minimum of 90% genome coverage and >200X median depth, but detection of the genomic variant was considered for final accuracy calculations. Because one erroneous SNP is unlikely to change genotyping conclusions in most instances, analysis was limited to overall variant detection using the default parameters for ease of implementation in the clinical laboratory. We also did not test the recommended 20 replicates to determine the LOD because this test is not intended to detect an analyte but the variations (genomic variant) in the analyte. Acceptable depth of coverage has been identified as 10X coverage of >90% of the genome. Low input of the RNA or lower reads will not meet these criteria. Therefore, false or undetermined variants are unlikely to be reported in low-input samples. Implementing a continuous performance measurement plan via an internal or external PT program is required to successfully integrate any test in the clinical laboratory (CAP Checklist 2021; https://www.cap.org). A set of reference SARS-CoV-2 variants is amenable to internal and external quality assurance testing. We assessed the entire workflow in preliminary internal PT by re-testing blind samples and inter-personnel reproducibility (details not shown).

WGS is a dynamic technology evolving rapidly; therefore, our validated pipeline is unlikely to remain static. Re-validation provision is crucial for the seamless and timely implementation of changes to wet-lab reagents or the analysis pipeline. We have introduced a provision for reagent verification at each lot change by re-testing samples in triplicate. Likewise, raw sequencing data will be re-analyzed with an updated analysis pipeline, and the accuracy of variant detection will be verified. DRAGEN COVID Lineage variant pipeline has been updated during the validation, and the variant identified by the two versions are in 100% concordance (data not shown). In general, WGS diagnosis reports are complex, and the format could pose challenges for the end-user. We adopted a simple format already used for SARS-CoV-2 diagnosis, and an updated report with the variant information will be issued if the reflex testing for variant detection is requested.

This study possesses certain limitations. First, only a limited number of WGS-based assays were included in the validation study based on the limited application of this test for clinical decision-making. Second, we could not establish this test’s clinical sensitivity and specificity because the clinical presentation of the patients infected with different variants is not distinct [30]. Moreover, the study could not acquire clinical samples of every lineage to demonstrate accuracy. However, CLIA and CAP regulations do not require validation of each mutation in the case of the mutation detection assay. For example, genomic mutation detection assays are commonly used in oncology. Likewise, we demonstrated the accuracy of the assay by testing three major variants. The vaccination status of the samples was not available to compare the application of the assay in the post-vaccination era. Although the vaccination status of the individual patient was not available, we tested >100 patient samples in the post-vaccination era to demonstrate that the test remains applicable to the vaccinated population. We could not try the test’s clinical utility because that would require enrolling the patients infected with different variants and administering variant-specific treatment.

Since their inception, most NGS-based testing has been limited to large medical centers, public health laboratories, or centralized genomics facilities with rather large infrastructures. The recent pandemic has accentuated the importance of de-centralized independent laboratories. For example, Advanta Genetics has served East Texas by testing > 500,000 SARS-CoV-2 samples. Thus, this validation can be used as guidelines for other small laboratories with NGS capacities if a need for SARS-CoV-2 variant detection arises. Although inevitable in the early stages, de-centralized NGS testing presents several challenges, such as high cost and turnaround time because of low volume testing. However, de-centralized and rapid testing for circulating SARS-CoV-2 variants may become crucial for clinical management and tracking the transmission at the local and regional levels. Sequencing cost in terms of dollars/gigabases has plummeted with high throughput instruments such as Illumina NovaSeq. However, such an instrument alone costs ∼ a million dollars. It would require batching of ∼30,000 samples to achieve the highest efficiency, which is not practical for independent laboratories, potentially leaving a gap in underserved communities.

This study introduces pre-pooling normalization to improve sequencing efficiency, which is crucial for smaller laboratories with low throughput sequencers. The emergence of more affordable sequencers such as Oxford Nanopore (Starting cost of $10,000) enriches the opportunity for de-centralized genomic testing if a variant with distinct clinical needs emerges or for any future pandemic. We have demonstrated that NGS services, including clinical testing, could be delivered locally with well-defined quality metrics at an affordable cost. Global NGS data aggregators that emerged from this pandemic have been helpful for analysis support needed for resource-limited laboratories (https://www.gisaid.org/collaborations/enabled-by-hcov-19-data-from-gisaid/), but sequencing infrastructure remains centralized mainly [31]. The local-delivery model would also be more responsive to the target clients’ needs and enhance the adoption of NGS across health care systems. We have demonstrated the application of this approach in the East Texas region and tracked the variant evolution throughout the pandemic. An alternate hybrid model has been proposed with complementary central and local services to balance the need for speed and investment [32]. The FDA Genome Tracker network for tracking foodborne pathogens and the Centers for Disease Control and Prevention (CDC) Advanced Molecular Detection (AMD) initiative for improving infectious disease surveillance are existing hybrid models in the United States [33, 34]. Notably, there are still significant challenges to implementing comprehensive WGS services locally [35, 36]. This study has established the performance specifications for NGS-based SARS-CoV-2 variant detection according to CAP and CLIA guidelines. We anticipate that the COVID-Seq LDT validation framework presented in this study, in synergy with increasingly accessible analysis support, will advance the localization of comprehensive NGS services in independent clinical laboratories. We have benchmarked quality assurance and quality control measures for implementing such testing and a simplified reporting format for end-users with limited NGS understanding. The study also affirmed the application of de-centralized NGS testing for clinical and public health applications with any resurgence of COVID-19 or the next infectious disease outbreak.

## Data Availability

All data produced in the present work are contained in the manuscript

## Author Contributions

**RC:** Conceived the study and assisted in the writing; **VT:** Performed the bioinformatics analysis and organized the data; **SA:** performed the experiments and organized the data; **EB:** performed the experiments; **RS*:** Designed the study, supervised experiments and wrote the manuscript. RC, VT, and SA contributed equally to this work and shared the first authorship.

### Ethical statement

The study was exempted by IRB (Institutional Review Board) of Advanta Genetics because only de-identified samples were used.

