## Supplementary Table 1 for "COVID Seq as Laboratory Developed Test (LDT) for diagnosis of SARS-CoV-2 Variants of Concern (VOC)"

SUPPLEMENTAL TABLE-1

**Data Availability**

GISAID Identifier: EPI_SET_20220715vh doi: 10.55876/gis8.220715vh

All genome sequences and associated metadata in this dataset are published in GISAID’s EpiCoV database. To view the contributors of each sequence with details such as accession number, Virus name, Collection date, Originating Lab and Submitting Lab, and the list of Authors, visit 10.55876/gis8.220715vh

**Data Snapshot**

EPI_SET_20220715vh is composed of 72 individual genome sequences.
The collection dates range from 2020-08-01 to 2022-09-25;
Data were collected in 1 country and territory;
All sequences in this dataset are compared to hCoV-19/Wuhan/WIV04/2019 (WIV04), the official reference sequence employed by GISAID (EPI_ISL_402124). Learn more at https://gisaid.org/WIV04.

Supplementary Table: 1- Demographic information of all 162 samples of SARS-COV-2, East Texas Region

| **Accession ID** | **GISAID Accession ID** | **Date of Collection** | **County** | **State** | **Run Date** | **PANGO Lineage** |
| --- | --- | --- | --- | --- | --- | --- |
| ACSQ1-12 | EPI_ISL_8719355 | Aug-20 | Smith | TX | 9/15/21 | B.1 |
| ACSQ1-8 | EPI_ISL_8729880 | Aug-20 | Smith | TX | 9/15/21 | B.1.243 |
| ACSQ1-13 | EPI_ISL_8729884 | Aug-20 | Smith | TX | 9/15/21 | B.1.234 |
| ACSQ1-14 | EPI_ISL_8729885 | Aug-20 | Smith | TX | 9/15/21 | B.1.126 |
| ACSQ1-15 | EPI_ISL_8729886 | Aug-20 | Smith | TX | 9/15/21 | B.1.602 |
| ACSQ1-16 | EPI_ISL_8729887 | Aug-20 | Smith | TX | 9/15/21 | B.1 |
| ACSQ1-17 | EPI_ISL_8729888 | Aug-20 | Smith | TX | 9/15/21 | B.1 |
| ACSQ1-18 | EPI_ISL_8729889 | Aug-20 | Smith | TX | 9/15/21 | B.1.564 |
| ACSQ1-19 | EPI_ISL_8729890 | Aug-20 | Smith | TX | 9/15/21 | B.1.617.2 |
| ACSQ1-20 | EPI_ISL_8729891 | Aug-20 | Smith | TX | 9/15/21 | AY.3 |
| ACSQ1-9 | EPI_ISL_8729881 | Aug-20 | Smith | TX | 9/15/21 | B.1.574 |
| ACSQ1-21 | EPI_ISL_8729892 | Aug-20 | Smith | TX | 9/15/21 | B.1.617.2 |
| ACSQ1-10 | EPI_ISL_8729882 | Aug-20 | Smith | TX | 9/15/21 | B.1.574 |
| ACSQ1-11 | EPI_ISL_8729883 | Aug-20 | Smith | TX | 9/15/21 | B.1.2 |
| ACSQ1-3 | EPI_ISL_8729875 | July-21 | Smith | TX | 9/15/21 | AY.3 |
| FT-SA99822 | EPI_ISL_3523224 | July-21 | SMITH | TX | 8/11/21 | AY.3 |
| FT-SA99824 | EPI_ISL_3523226 | July-21 | SMITH | TX | 8/11/21 | AY.3 |
| FT-SA99827 | EPI_ISL_3523229 | July-21 | SMITH | TX | 8/11/21 | AY.3 |
| FT-SA99828 | EPI_ISL_3523230 | July-21 | BRAZOS | TX | 8/11/21 | B.1.617.2 |
| FT-SA99833 | EPI_ISL_3523235 | July-21 | SMITH | TX | 8/11/21 | AY.3 |
| ACSQ1-5 | EPI_ISL_8729877 | July-21 | Smith | TX | 9/15/21 | AY.3 |
| FT-SA99772 | EPI_ISL_3523183 | July-21 | TARRANT | TX | 8/11/21 | B.1.617.2 |
| FT-SA99802 | EPI_ISL_3523208 | July-21 | VAN ZANDT | TX | 8/11/21 | AY.3 |
| FT-SA99820 | EPI_ISL_3523222 | July-21 | SMITH | TX | 8/11/21 | AY.3 |
| FT-SA99821 | EPI_ISL_3523223 | July-21 | SMITH | TX | 8/11/21 | AY.3 |
| FT-SA99823 | EPI_ISL_3523225 | July-21 | SMITH | TX | 8/11/21 | AY.3 |
| FT-SA99825 | EPI_ISL_3523227 | July-21 | SMITH | TX | 8/11/21 | AY.3 |
| FT-SA99826 | EPI_ISL_3523228 | July-21 | SMITH | TX | 8/11/21 | AY.3 |
| FT-SA99829 | EPI_ISL_3523231 | July-21 | SMITH | TX | 8/11/21 | AY.3 |
| FT-SA99830 | EPI_ISL_3523232 | July-21 | SMITH | TX | 8/11/21 | B.1.617.2 |
| FT-SA99832 | EPI_ISL_3523234 | July-21 | JOHNSON | TX | 8/11/21 | AY.3 |
| FT-SA99852 | EPI_ISL_3523252 | July-21 | SMITH | TX | 8/11/21 | AY.3 |
| FT-SA99856 | EPI_ISL_3523256 | July-21 | SMITH | TX | 8/11/21 | B.1.617.2 |
| FT-SA99861 | EPI_ISL_3523261 | July-21 | SMITH | TX | 8/11/21 | B.1.617.2 |
| FT-SA99863 | EPI_ISL_3523262 | July-21 | GREGG | TX | 8/11/21 | B.1.617.2 |
| ACSQ1-7 | EPI_ISL_8729879 | July-21 | Smith | TX | 9/15/21 | AY.3 |
| ACSQ1-2 | EPI_ISL_8729874 | July-21 | Smith | TX | 9/15/21 | AY.25 |
| FT-SA99774 | EPI_ISL_3523185 | July-21 | HIDALGO | TX | 8/11/21 | B.1.617.2 |
| FT-SA99785 | EPI_ISL_3523194 | July-21 | SMITH | TX | 8/11/21 | AY.3 |
| FT-SA99790 | EPI_ISL_3523198 | July-21 | SMITH | TX | 8/11/21 | AY.3 |
| FT-SA99791 | EPI_ISL_3523199 | July-21 | SMITH | TX | 8/11/21 | AY.3 |
| FT-SA99792 | EPI_ISL_3523200 | July-21 | HARRIS | TX | 8/11/21 | B.1.617.2 |
| FT-SA99793 | EPI_ISL_3523201 | July-21 | SMITH | TX | 8/11/21 | AY.3 |
| FT-SA99795 | EPI_ISL_3523202 | July-21 | SMITH | TX | 8/11/21 | AY.3 |
| FT-SA99796 | EPI_ISL_3523203 | July-21 | JOHNSON | TX | 8/11/21 | B.1.617.2 |
| FT-SA99803 | EPI_ISL_3523209 | July-21 | SMITH | TX | 8/11/21 | AY.3 |
| FT-SA99804 | EPI_ISL_3523210 | July-21 | HOOD | TX | 8/11/21 | B.1.617.2 |
| FT-SA99831 | EPI_ISL_3523233 | July-21 | TARRANT | TX | 8/11/21 | B.1.617.2 |
| FT-SA99835 | EPI_ISL_3523237 | July-21 | SMITH | TX | 8/11/21 | AY.3 |
| FT-SA99840 | EPI_ISL_3523241 | July-21 | SMITH | TX | 8/11/21 | AY.3 |
| FT-SA99841 | EPI_ISL_3523242 | July-21 | SMITH | TX | 8/11/21 | AY.3 |
| FT-SA99850 | EPI_ISL_3523250 | July-21 | SMITH | TX | 8/11/21 | AY.3 |
| FT-SA99858 | EPI_ISL_3523258 | July-21 | SMITH | TX | 8/11/21 | AY.3 |
| FT-SA99859 | EPI_ISL_3523259 | July-21 | SMITH | TX | 8/11/21 | AY.3 |
| FT-SA99860 | EPI_ISL_3523260 | July-21 | SMITH | TX | 8/11/21 | B.1.617.2 |
| FT-SA99865 | EPI_ISL_3523264 | July-21 | SMITH | TX | 8/11/21 | AY.3 |
| ACSQ1-4 | EPI_ISL_8729876 | July-21 | Smith | TX | 9/15/21 | AY.3 |
| FT-SA99770 | EPI_ISL_3523181 | July-21 | SMITH | TX | 8/11/21 | B.1.617.2 |
| FT-SA99771 | EPI_ISL_3523182 | July-21 | SMITH | TX | 8/11/21 | AY.3 |
| FT-SA99773 | EPI_ISL_3523184 | July-21 | BRAZOS | TX | 8/11/21 | B.1.617.2 |
| FT-SA99775 | EPI_ISL_3523186 | July-21 | SMITH | TX | 8/11/21 | AY.3 |
| FT-SA99776 | EPI_ISL_3523187 | July-21 | HARRISON | TX | 8/11/21 | AY.3 |
| FT-SA99777 | EPI_ISL_3523188 | July-21 | SMITH | TX | 8/11/21 | AY.3 |
| FT-SA99780 | EPI_ISL_3523189 | July-21 | SMITH | TX | 8/11/21 | B.1.617.2 |
| FT-SA99781 | EPI_ISL_3523190 | July-21 | SMITH | TX | 8/11/21 | B.1.617.2 |
| FT-SA99782 | EPI_ISL_3523191 | July-21 | SMITH | TX | 8/11/21 | AY.3 |
| FT-SA99783 | EPI_ISL_3523192 | July-21 | SMITH | TX | 8/11/21 | AY.3 |
| FT-SA99784 | EPI_ISL_3523193 | July-21 | SMITH | TX | 8/11/21 | AY.3 |
| FT-SA99786 | EPI_ISL_3523195 | July-21 | SMITH | TX | 8/11/21 | AY.3 |
| FT-SA99788 | EPI_ISL_3523196 | July-21 | SMITH | TX | 8/11/21 | AY.3 |
| FT-SA99789 | EPI_ISL_3523197 | July-21 | SMITH | TX | 8/11/21 | AY.3 |
| FT-SA99797 | EPI_ISL_3523204 | July-21 | SMITH | TX | 8/11/21 | AY.3 |
| FT-SA99799 | EPI_ISL_3523205 | July-21 | SMITH | TX | 8/11/21 | AY.3 |
| FT-SA99800 | EPI_ISL_3523206 | July-21 | SMITH | TX | 8/11/21 | B.1.617.2 |
| FT-SA99801 | EPI_ISL_3523207 | July-21 | HENDERSON | TX | 8/11/21 | B.1.617.2 |
| FT-SA99805 | EPI_ISL_3523211 | July-21 | SMITH | TX | 8/11/21 | B.1.617.2 |
| FT-SA99809 | EPI_ISL_3523214 | July-21 | SMITH | TX | 8/11/21 | B.1.617.2 |
| ACSQ1-6 | EPI_ISL_8729878 | July-21 | Smith | TX | 9/15/21 | B.1.617.2 |
| FT-SA99807 | EPI_ISL_3523212 | July-21 | SMITH | TX | 8/11/21 | B.1.617.2 |
| FT-SA99808 | EPI_ISL_3523213 | July-21 | SMITH | TX | 8/11/21 | B.1.617.2 |
| FT-SA99813 | EPI_ISL_3523215 | July-21 | JOHNSON | TX | 8/11/21 | B.1.617.2 |
| FT-SA99815 | EPI_ISL_3523217 | July-21 | SMITH | TX | 8/11/21 | AY.3 |
| FT-SA99816 | EPI_ISL_3523218 | July-21 | SMITH | TX | 8/11/21 | AY.3 |
| FT-SA99817 | EPI_ISL_3523219 | July-21 | SMITH | TX | 8/11/21 | AY.3 |
| FT-SA99818 | EPI_ISL_3523220 | July-21 | SMITH | TX | 8/11/21 | AY.3 |
| FT-SA99819 | EPI_ISL_3523221 | July-21 | HENDERSON | TX | 8/11/21 | B.1.617.2 |
| FT-SA99814 | EPI_ISL_3523216 | July-21 | SMITH | TX | 8/11/21 | AY.3 |
| ACSQ4-23 | EPI_ISL_8629675 | Dec-21 | Passaic | NJ | 12/26/21 | B.1.1.529 |
| ACSQ4-22 | EPI_ISL_8629674 | Dec-21 | SMITH | TX | 12/26/21 | B.1.1.529 |
| ACSQ4-21 | EPI_ISL_8629673 | Dec-21 | Tarrant | TX | 12/26/21 | AY.111 |
| ACSQ4-19 | EPI_ISL_8629671 | Dec-21 | SMITH | TX | 12/26/21 | BA.1 |
| ACSQ4-1 | EPI_ISL_8629666 | Dec-21 | Tarrant | TX | 12/26/21 | B.1.1.529 |
| ACSQ4-58 | EPI_ISL_8428058 | Dec-21 | SMITH | TX | 12/26/21 | BA.1 |
| ACSQ4-4 | EPI_ISL_8629668 | Dec-21 | Smith | TX | 12/26/21 | AY.103 |
| ACSQ4-18 | EPI_ISL_8629670 | Dec-21 | Navarro | TX | 12/26/21 | B.1.1.529 |
| ACSQ4-2 | EPI_ISL_8629667 | Dec-21 | Navarro | TX | 12/26/21 | B.1.1.529 |
| ACSQ4-20 | EPI_ISL_8629672 | Dec-21 | Navarro | TX | 12/26/21 | AY.3 |
| ACSQ4-24 | EPI_ISL_8629676 | Dec-21 | Passaic | NJ | 12/26/21 | B.1.1.529 |
| ACSQ4-17 | EPI_ISL_8629669 | Dec-21 | SMITH | TX | 12/26/21 | B.1.1.529 |
| ACSQ7-1 | EPI_ISL_13655593 | April-22 | Van Zandt | TX | 6/24/22 | BA.2 |
| ACSQ7-2 | EPI_ISL_13655594 | April-22 | Smith | TX | 6/24/22 | BA.3 |
| ACSQ7-11 | EPI_ISL_13655603 | April-22 | Smith | TX | 6/24/22 | BA.2.3 |
| ACSQ7-17 | EPI_ISL_13655607 | May-22 | Brazos | TX | 6/24/22 | BA.5 |
| ACSQ7-15 | EPI_ISL_13671513 | May-22 | Brazos | TX | 6/24/22 | BA.2.9 |
| ACSQ7-3 | EPI_ISL_13655595 | May-22 | Smith | TX | 6/24/22 | BA.2.12.1 |
| ACSQ7-12 | EPI_ISL_13655604 | May-22 | Brazos | TX | 6/24/22 | BA.2.3 |
| ACSQ7-16 | EPI_ISL_13655606 | May-22 | Van Zandt | TX | 6/24/22 | BA.2.9 |
| ACSQ7-5 | EPI_ISL_13655597 | May-22 | Smith | TX | 6/24/22 | BA.2.12.1 |
| ACSQ7-4 | EPI_ISL_13655596 | May-22 | Smith | TX | 6/24/22 | BA.2.12.1 |
| ACSQ7-7 | EPI_ISL_13655599 | May-22 | Tarrant | TX | 6/24/22 | BA.2.12.1 |
| ACSQ7-6 | EPI_ISL_13655598 | May-22 | Smith | TX | 6/24/22 | BA.2.12.1 |
| ACSQ8-1 | EPI_ISL_13762660 | May-22 | Gregg | TX | 7/9/22 | BA.2.12.1 |
| ACSQ7-13 | EPI_ISL_13671512 | May-22 | Smith | TX | 6/24/22 | BA.2.37 |
| ACSQ7-8 | EPI_ISL_13655600 | May-22 | Henderson | TX | 6/24/22 | BA.2.12.1 |
| ACSQ7-14 | EPI_ISL_13655605 | May-22 | Johnson | TX | 6/24/22 | BA.2.37 |
| ACSQ8-2 | EPI_ISL_13819250 | Jun-22 | Gregg | TX | 7/9/22 | BA.2.12.1 |
| ACSQ7-10 | EPI_ISL_13655602 | Jun-22 | Smith | TX | 6/24/22 | BA.2.12.1 |
| ACSQ7-9 | EPI_ISL_13655601 | Jun-22 | Smith | TX | 6/24/22 | BA.2.12.1 |
| ACSQ8-3 | EPI_ISL_13819251 | Jun-22 | Gregg | TX | 7/9/22 | BA.2.12.1 |
| ACSQ8-4 | EPI_ISL_13762661 | Jun-22 | Gregg | TX | 7/9/22 | BA.2.12.1 |
| ACSQ8-5 | EPI_ISL_13762662 | Jun-22 | Gregg | TX | 7/9/22 | BA.2.12.1 |
| ACSQ8-7 | EPI_ISL_13762664 | Jun-22 | Gregg | TX | 7/9/22 | BA.5.2.1 |
| ACSQ8-6 | EPI_ISL_13762663 | Jun-22 | Gregg | TX | 7/9/22 | BA.5.5 |
| ACSQ8-8 | EPI_ISL_13762665 | Jun-22 | Gregg | TX | 7/9/22 | BA.2.12.1 |
| ACSQ8-9 | EPI_ISL_13762666 | Jun-22 | Gregg | TX | 7/9/22 | BA.2.12.1 |
| ACSQ8-10 | EPI_ISL_13762667 | Jun-22 | Gregg | TX | 7/9/22 | BA.4.1 |
| ACSQ8-11 | EPI_ISL_13762668 | Jun-22 | Gregg | TX | 7/9/22 | BA.2.12.1 |
| ACSQ8-12 | EPI_ISL_13819252 | Jun-22 | Gregg | TX | 7/9/22 | BA.2.12.1 |
| ACSQ8-14 | EPI_ISL_13819253 | Jun-22 | Gregg | TX | 7/9/22 | BA.2.12.1 |
| ACSQ8-13 | EPI_ISL_13762669 | July-22 | Gregg | TX | 7/9/22 | BA.2.12.1 |
| ACSQ8-15 | EPI_ISL_13762670 | July-22 | Gregg | TX | 7/9/22 | BA.2.12.1 |
| ACSQ8-16 | EPI_ISL_13762671 | July-22 | Gregg | TX | 7/9/22 | BA.2.12.1 |
| ACSQ8-18 | EPI_ISL_13762673 | July-22 | Gregg | TX | 7/9/22 | BA.2.12.1 |
| ACSQ8-17 | EPI_ISL_13762672 | July-22 | Gregg | TX | 7/9/22 | BA.2.12.1 |
| ACSQ8-19 | EPI_ISL_13762674 | July-22 | Gregg | TX | 7/9/22 | BA.5.2.1 |
| ACSQ8-22 | EPI_ISL_13762677 | July-22 | Gregg | TX | 7/9/22 | BA.2.12.1 |
| ACSQ8-23 | EPI_ISL_13762678 | July-22 | Gregg | TX | 7/9/22 | BA.2.12.1 |
| ACSQ8-20 | EPI_ISL_13762675 | July-22 | Gregg | TX | 7/9/22 | BA.5.2.1 |
| ACSQ8-21 | EPI_ISL_13762676 | July-22 | Gregg | TX | 7/9/22 | BA.2.12.1 |
| ACSQ9-15 | EPI_ISL_15169383 | Sep-22 | Smith | TX | 9/26/22 | BA.5.2 |
| ACSQ9-24 | EPI_ISL_15169391 | Sep-22 | Henderson | TX | 9/26/22 | BA.5.2.1 |
| ACSQ9-23 | EPI_ISL_15169390 | Sep-22 | Smith | TX | 9/26/22 | BA.2.3 |
| ACSQ9-22 | EPI_ISL_15169389 | Sep-22 | Anderson | TX | 9/26/22 | BA.5.5 |
| ACSQ9-17 | EPI_ISL_15169385 | Sep-22 | Tarrant | TX | 9/26/22 | BA.5.6 |
| ACSQ9-18 | EPI_ISL_15169386 | Sep-22 | Henderson | TX | 9/26/22 | BA.5.1.6 |
| ACSQ9-21 | EPI_ISL_15169388 | Sep-22 | Shelby | TX | 9/26/22 | BA.5.2.1 |
| ACSQ9-14 | EPI_ISL_15169382 | Sep-22 | Tarrant | TX | 9/26/22 | BA.5.2 |
| ACSQ9-20 | EPI_ISL_15169387 | Sep-22 | Nacogdoches | TX | 9/26/22 | BA.5.2 |
| ACSQ9-13 | EPI_ISL_15169381 | Sep-22 | Smith | TX | 9/26/22 | BA.4.6 |
| ACSQ9-12 | EPI_ISL_15169380 | Sep-22 | Smith | TX | 9/26/22 | BA.4.6 |
| ACSQ9-7 | EPI_ISL_15169375 | Sep-22 | Smith | TX | 9/26/22 | BE.1.1 |
| ACSQ9-16 | EPI_ISL_15169384 | Sep-22 | Anderson | TX | 9/26/22 | BA.5.5 |
| ACSQ9-8A | EPI_ISL_15169376 | Sep-22 | Denton | TX | 9/26/22 | BA.5.2 |
| ACSQ9-9 | EPI_ISL_15169377 | Sep-22 | Smith | TX | 9/26/22 | BE.1.1 |
| ACSQ9-11 | EPI_ISL_15169379 | Sep-22 | Brazos | TX | 9/26/22 | BA.4.6 |
| ACSQ9-10 | EPI_ISL_15169378 | Sep-22 | Gregg | TX | 9/26/22 | BA.5.2 |
| ACSQ9-3 | EPI_ISL_15169371 | Sep-22 | Johnson | TX | 9/26/22 | BA.5.1.1 |
| ACSQ9-5 | EPI_ISL_15169373 | Sep-22 | Tarrant | TX | 9/26/22 | BA.5.1 |
| ACSQ9-4A | EPI_ISL_15169372 | Sep-22 | Tarrant | TX | 9/26/22 | BA.5.2.1 |
| ACSQ9-2 | EPI_ISL_15169370 | Sep-22 | Smith | TX | 9/26/22 | BA.5.2 |
| ACSQ9-6 | EPI_ISL_15169374 | Sep-22 | Henderson | TX | 9/26/22 | BA.5.1 |
| ACSQ9-1A | EPI_ISL_15169369 | Sep-22 | Smith | TX | 9/26/22 | BA.4.6 |
