## Supplementary Table 2 for "COVID Seq as Laboratory Developed Test (LDT) for diagnosis of SARS-CoV-2 Variants of Concern (VOC)"

Supplementary Table: 3- Inter-Sequencing run precision: Variant identification concordance across three different library preparation and sequencing runs

| **Accession** | **ACSQ-2 (Rep-1)** | | | | **ACSQ-3 (Rep-2)** | | | | **ACSQ-4 (Rep-3)** | | | | **Concordance** |
| --- | --- | --- | --- | --- | --- | --- | --- | --- | --- | --- | --- | --- | --- |
|  | **Median Coverage-R1** | **Coverage >= 30x-R1** | **Pango  Lineage** | **WHO label** | **Median Coverage-R2** | **Coverage >= 30xR-2** | **Pango  Lineage** | **WHO label** | **Median Coverage-3** | **Coverage >= 30x-R3** | **Pango  Lineage** | **WHO label** |  |
| ACSQ1-3 | 345 | 86.06% | AY.3 | Delta (B.1.617.2-like) | 120 | 80.09% | AY.3 | Delta (B.1.617.2-like) | 149 | 84.77% | AY.82 | Delta (B.1.617.2-like) | YES |
| ACSQ1-7 | 2289 | 99.78% | AY.25 | Delta (B.1.617.2-like) | 364 | 99.33% | AY.25 | Delta (B.1.617.2-like) | 444 | 98.70% | AY.25 | Delta (B.1.617.2-like) | YES |
| ACSQ1-2 | 797 | 99.01% | AY.3 | Delta (B.1.617.2-like) | 338 | 98.92% | AY.3 | Delta (B.1.617.2-like) | 418 | 98.34% | AY.3 | Delta (B.1.617.2-like) | YES |
| ACSQ2-14 | 547 | 92.60% | AY.3 | Delta (B.1.617.2-like) | 226 | 92.85% | AY.3 | Delta (B.1.617.2-like) | 241 | 94.21% | AY.3 | Delta (B.1.617.2-like) | YES |
| ACSQ2-20 | 757 | 99.59% | AY.39.1 | Delta (B.1.617.2-like) | 310 | 99.39% | AY.39.1 | Delta (B.1.617.2-like) | 383 | 99.01% | AY.39.1 | Delta (B.1.617.2-like) | YES |
| ACSQ2-17 | 633 | 95.39% | AY.103 | Delta (B.1.617.2-like) | 265 | 95.65% | AY.103 | Delta (B.1.617.2-like) | 298 | 97.21% | AY.103 | Delta (B.1.617.2-like) | YES |
| ACSQ4-1 | 1286 | 98.32% | B.1.1.529 | Omicron (B.1.1.529-like) | 1363 | 99.15% | B.1.1.529 | Omicron (B.1.1.529-like) | 1209 | 97.54% | B.1.1.529 | Omicron (B.1.1.529-like) | YES |
| ACSQ4-85 | 1131 | 96.64% | B.1.1.529 | Omicron (B.1.1.529-like) | 1201 | 97.10% | B.1.1.529 | Omicron (B.1.1.529-like) | 1308 | 96.61% | B.1.1.529 | Omicron (B.1.1.529-like) | YES |
| ACSQ4-2 | 867 | 92.42% | B.1.1.529 | Omicron (B.1.1.529-like) | 1130 | 94.96% | B.1.1.529 | Omicron (B.1.1.529-like) | 900 | 95.46% | B.1.1.529 | Omicron (B.1.1.529-like) | YES |
