## Supplementary Table 3 for "COVID Seq as Laboratory Developed Test (LDT) for diagnosis of SARS-CoV-2 Variants of Concern (VOC)"

Supplementary Table: 2- Intra-Sequencing run precision: Variant identification concordance among the triplicate testing of the same samples

| **Accession** | **(Rep-1)** | | | | **(Rep-2)** | | | | **(Rep-3)** | | | | **Concordance** |
| --- | --- | --- | --- | --- | --- | --- | --- | --- | --- | --- | --- | --- | --- |
|  | **Median Coverage** | **Coverage >= 30x** | **Pango  Lineage** | **WHO label** | **Median Coverage** | **Coverage >= 30x** | **Pango  Lineage** | **WHO label** | **Median Coverage** | **Coverage >= 30x** | **Pango  Lineage** | **WHO label** |  |
| AACSQ1-3 | 345 | 86.06% | AY.3 | Delta (B.1.617.2-like) | 302 | 86.55% | AY.3 | Delta (B.1.617.2-like) | 309 | 85.61% | AY.3 | Delta (B.1.617.2-like) | YES |
| AACSQ1-3* | 120 | 80.09% | AY.3 | Delta (B.1.617.2-like) | 108 | 81.39% | None |  | 107 | 79.47% | AY.3 | Delta (B.1.617.2-like) | Inconclusive |
| AACSQ1-3* | 149 | 84.77% | AY.82 | Delta (B.1.617.2-like) | 132 | 84.38% | None |  | 85 | 74.01% | AY.3 | Delta (B.1.617.2-like) | Inconclusive |
| ACSQ1-7 | 2289 | 99.78% | AY.25 | Delta (B.1.617.2-like) | 684 | 99.19% | AY.25 | Delta (B.1.617.2-like) | 773 | 99.39% | AY.25 | Delta (B.1.617.2-like) | YES |
| ACSQ1-7 | 364 | 99.33% | AY.25 | Delta (B.1.617.2-like) | 289 | 99.14% | AY.25 | Delta (B.1.617.2-like) | 371 | 98.61% | AY.25 | Delta (B.1.617.2-like) | YES |
| ACSQ1-7 | 444 | 98.70% | AY.25 | Delta (B.1.617.2-like) | 303 | 98.43% | AY.25 | Delta (B.1.617.2-like) | 435 | 98.64% | AY.25 | Delta (B.1.617.2-like) | YES |
| ACSQ1-2 | 338 | 98.92% | AY.3 | Delta (B.1.617.2-like) | 349 | 98.97% | AY.3 | Delta (B.1.617.2-like) | 316 | 0.9892 | AY.3 | Delta (B.1.617.2-like) | YES |
| ACSQ1-2 | 418 | 98.34% | AY.3 | Delta (B.1.617.2-like) | 427 | 98.45% | AY.3 | Delta (B.1.617.2-like) | 336 | 98.30% | AY.3 | Delta (B.1.617.2-like) | YES |
| ACSQ2-14 | 547 | 92.60% | AY.3 | Delta (B.1.617.2-like) | 446 | 92.25% | AY.3 | Delta (B.1.617.2-like) | 560 | 93.01% | AY.3 | Delta (B.1.617.2-like) | YES |
| ACSQ2-14 | 226 | 92.85% | AY.3 | Delta (B.1.617.2-like) | 165 | 91.83% | AY.3 | Delta (B.1.617.2-like) | 224 | 93.91% | AY.3 | Delta (B.1.617.2-like) | YES |
| ACSQ2-14 | 241 | 94.21% | AY.3 | Delta (B.1.617.2-like) | 227 | 94.18% | AY.3 | Delta (B.1.617.2-like) | 242 | 94.38% | AY.3 | Delta (B.1.617.2-like) | YES |
| ACSQ2-20 | 757 | 99.59% | AY.39.1 | Delta (B.1.617.2-like) | 706 | 99.59% | AY.39.1 | Delta (B.1.617.2-like) | 786 | 99.59% | AY.39.1 | Delta (B.1.617.2-like) | YES |
| ACSQ2-20 | 310 | 99.39% | AY.39.1 | Delta (B.1.617.2-like) | 295 | 99.37% | AY.39.1 | Delta (B.1.617.2-like) | 330 | 99.70% | AY.39.1 | Delta (B.1.617.2-like) | YES |
| ACSQ2-20 | 383 | 99.01% | AY.39.1 | Delta (B.1.617.2-like) | 371 | 98.89% | AY.39.1 | Delta (B.1.617.2-like) | 410 | 98.92% | AY.39.1 | Delta (B.1.617.2-like) | YES |
| ACSQ2-17 | 633 | 95.39% | AY.103 | Delta (B.1.617.2-like) | 583 | 95.43% | AY.103 | Delta (B.1.617.2-like) | 595 | 95.20% | AY.103 | Delta (B.1.617.2-like) | YES |
| ACSQ2-17 | 265 | 95.65% | AY.103 | Delta (B.1.617.2-like) | 237 | 94.94% | AY.103 | Delta (B.1.617.2-like) | 263 | 95.17% | AY.103 | Delta (B.1.617.2-like) | YES |
| ACSQ2-17 | 298 | 97.21% | AY.103 | Delta (B.1.617.2-like) | 275 | 96.96% | AY.103 | Delta (B.1.617.2-like) | 247 | 96.38% | AY.103 | Delta (B.1.617.2-like) | YES |
| ACSQ2-AB | 1781 | 98.24% | BA.5.2 | Omicron (BA.5-like) | 1249 | 96.34% | BA.5.2 | Omicron (BA.5-like) | 2050 | 97.95% | BA.5.2 | Omicron (BA.5-like) | YES |

*Variant detection was inconclusive because of the low coverage
